# Monitoring Disease Transmissibility of 2019 Novel Coronavirus Disease in Zhejiang, China

**DOI:** 10.1101/2020.03.02.20028704

**Authors:** Ka Chun Chong, Wei Cheng, Shi Zhao, Feng Ling, Kirran N. Mohammad, Maggie Haitian Wang, Benny Chung Ying Zee, Lei Wei, Xi Xiong, Hengyan Liu, Jiangxuan Wang, Enfu Chen

## Abstract

We monitored the transmissibility of 2019 novel coronavirus disease in Zhejiang accounting the transmissions from imported cases. Even though Zhejiang is one of the worst-affected provinces, an interruption of disease transmission (i.e. instantaneous reproduction numbers <1) was observed in early/mid-February after an early social-distancing response to the outbreak.

## Text

In December 2019, a novel strain of coronavirus emerged and caused an outbreak in Wuhan, Hubei province, China *(1)*. The disease responsible for the outbreak has been officially named by the World Health Organization (WHO) as COVID-19. Common clinical manifestation of COVID-19 includes fever, fatigue, dry cough, dyspnea and muscle ache *(2)*.

The current global scale health crisis began on 31 December 2019, the day WHO was notified of several unusual cases of pneumonia in Wuhan *(3)*. Despite the suspected source of transmission (Huanan Seafood Wholesale Market) was shut down the next day, number of cases continued to expand at an alarming rate *(3)*. On 9 January 2020, the first related death was recorded in Wuhan *(4)*; meanwhile, the disease spread outside Hubei as people travel around and outside the country. As of 22 February 2020, the epidemic registered 76,392 cases with 2,348 deaths in mainland China and spread to 28 countries that reported a total of 1,402 cases *(5)*.

To halt the spread of COVID-19, the Chinese government imposed a complete lockdown in Wuhan and other cities in Hubei province on 23 January 2020 aiming to quarantine the epicenter of the outbreak *(6)*. Meanwhile, a number of provinces/cities in China have imposed restrictions on entry by travelers from Wuhan in response to the epidemic.

Owing to the frequent travel connections between Wuhan and Zhejiang (an eastern province with 57 million residents), Zhejiang is the third worst-affected province with 1,205 cases confirmed to date, and the first province to declare the highest provincial level public health emergency in response to the outbreak (23 January 2020); officials closed non-essential public venues, banned funerals and weddings, and ordered suspension of work. Arrivals with travel history to other provinces within last 14 days shall be quarantined. Starting from 1 February to 6 February 2020, all the 11 cities in Zhejiang imposed lockdown for partial districts (Zhoushan, Jiaxing, Quzhou, Shaoxing, Lishui, Jinhua, and Huzhou) and a complete lockdown for all districts (Hangzhou, Ningbo, Wenzhou, and Taizhou). During the lockdown period, only one person per household is allowed to leave home once every two days for supplies *(7)*. In this study, we estimated the instantaneous reproduction number (*R*_*t*_) of COVID-19 in Zhejiang to monitor the impact of control measures on disease transmissibility over time.

### The Study

We analyzed the data of confirmed cases, including both local and imported cases from 26 December 2019 to 25 February 2020, from all cities in Zhejiang. Confirmed cases were tested positive for COVID-19, either through real-time reverse-transcription– polymerase-chain-reaction (RT-PCR) assay or genetic sequencing, in accordance with the WHO guidelines. Cases with travel history to other provinces (with ≥1 case reported) 14 days prior to symptoms onset are defined as import cases. Epidemiological data of confirmed cases were recorded to the database of Zhejiang Provincial Center for Disease Control and Prevention. Data were reviewed independently by at least two staffs to ensure data accuracy.

*R*_*t*_ is defined as the average number of secondary cases generated by a primary case at time *t* in a population and it is a time-varying measure of disease transmissibility when intervention measures are in place for outbreak control. If *R*_*t*_ is below unity, a disease transmission is unlikely to be sustained and the outbreak shall be under control. *R*_*t*_ can thus be used to monitor the disease evolution in a population over time *(8)*. In this study, we employed two different methods to estimate *R*_*t*_: 1. Wallinga and Teunis approach (M1) *(9,10)* and 2. susceptible-exposed-infectious-recovered (SEIR) model-based approach (M2), controlling for transmission arising from imported cases. In M1, we assumed serial interval follows a lognormal distribution with mean 7.5 days and standard deviation 3.4 days *(11)*, whereas in M2, we assumed the mean latent duration and mean infectious duration to be 5.2 *(11)* and 2.3 days respectively. Using the time series data of local and imported cases, we formulated the likelihoods and solved the model parameter *R*_*t*_ (additional details in the Appendix).

The first imported case of COVID-19 had illness onset on 1 January 2020. As importation of infections continued, the first local case had symptoms onset on 10 January 2020 (Figure 1). Over the study period, *R*_*t*_ attained peak values of 2.08 (95% confidence interval [CI]: 1.49 to 1.72) and 1.88 (95% CI: 1.38 to 2.41) on 16 January 2020 for method M1 and M2 respectively (Figure 2), and a majority of cases (60%) was still the imported cases. The *R*_*t*_s remained stable and kept above 1.40 until 28 January 2020, five days after the declaration of the highest-level public health emergency. Although the number of local cases had gradually surpassed that of imported cases, the *R*_*t*_s decreased over time. After the lockdown of cities, the upper bounds of CI of *R*_*t*_ for M1 and M2 were below the threshold of unity on 6 February 2020 and 12 February 2020 respectively (M1: 0.91 [95% CI: 0.87 to 0.96] and M2: 0.94 [95% CI: 0.90 to 0.98]). The *R*_*t*_s were to be 0.63 (95% CI: 0.60 to 0.65) for M1 and 0.78 (95% CI: 0.75 to 0.82) for M2 on 20 February 2020, the day the last cases were reported to date. Overall, the estimates from two different methods were consistent and showed a similar trend of *R*_*t*_. An assumption of gamma-distributed serial interval was tested and the results were consistent (Appendix Figure).

**Figure 1.**
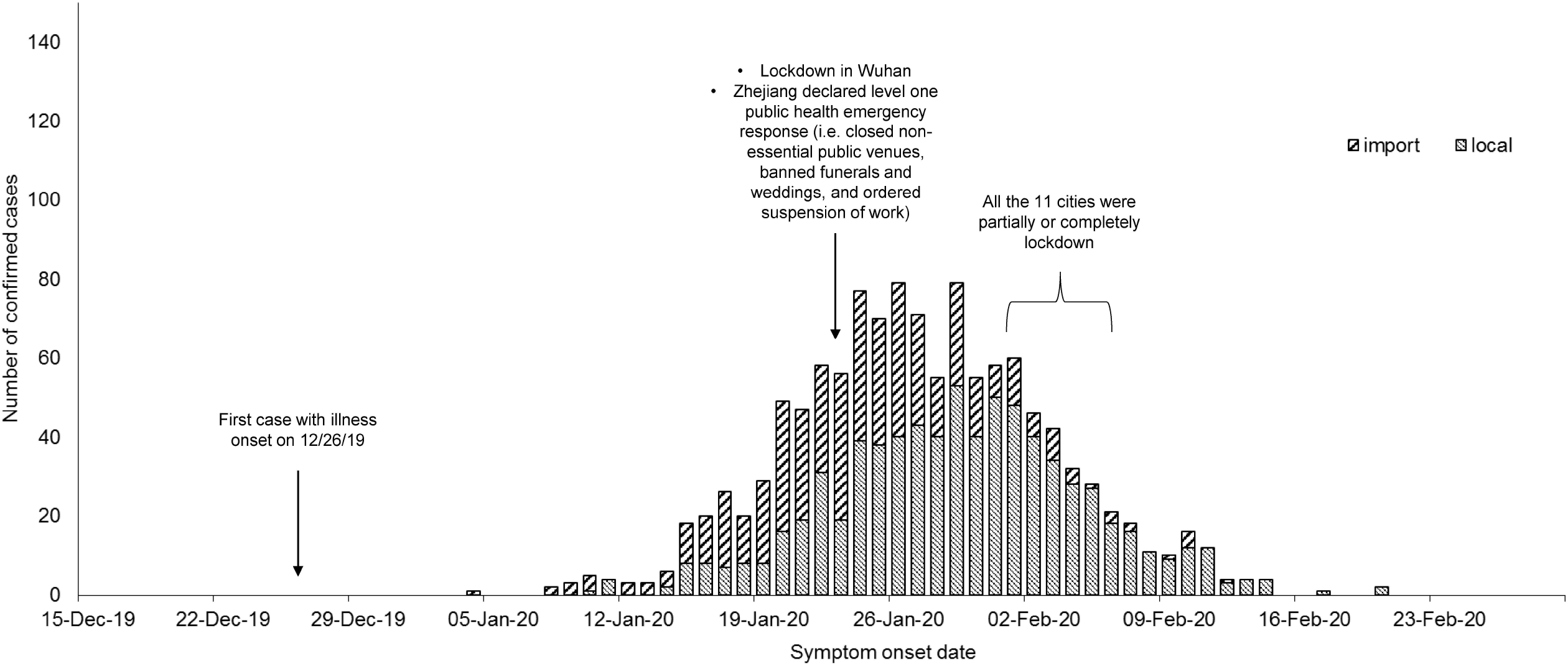
Number of imported cases and local cases of COVID-19 in Zhejiang against symptom-onset date

**Figure 2.**
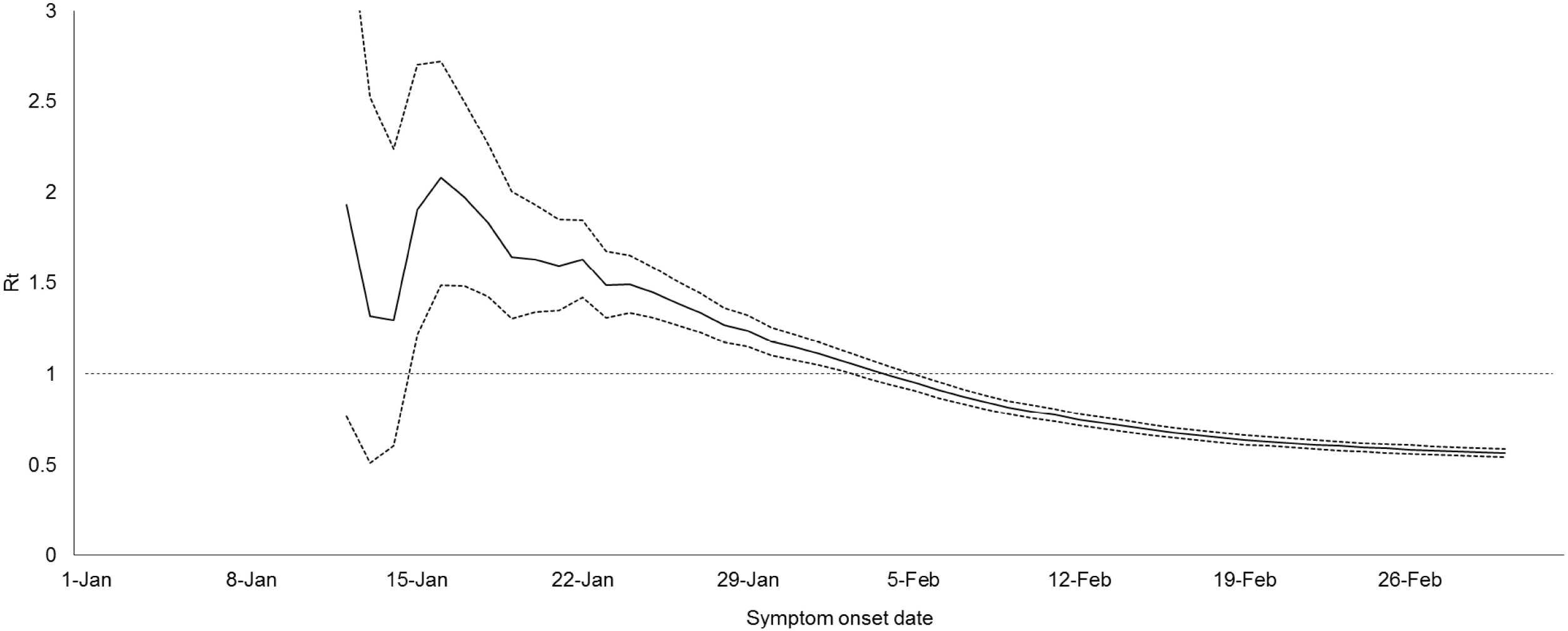
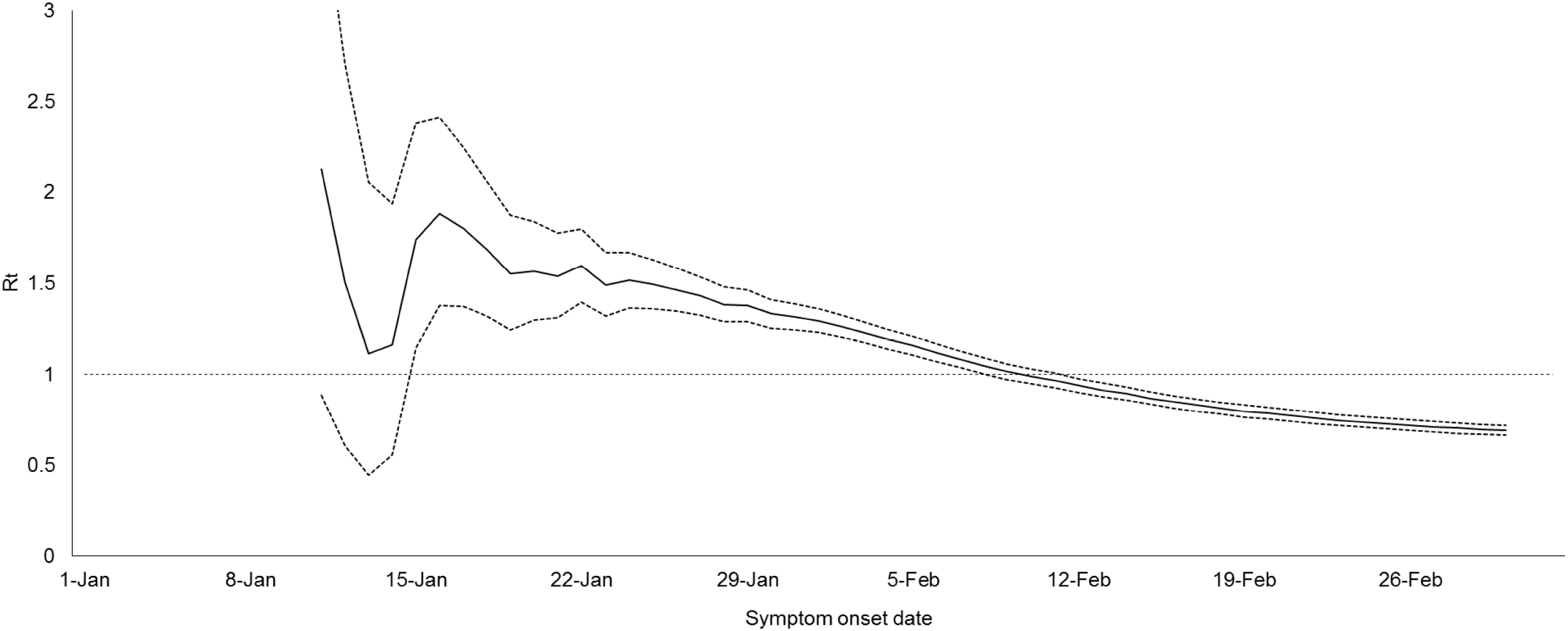
Estimated instantaneous reproduction number (*R*_*t*_) (solid line) and 95% confidence intervals (dotted lines) using (A) Wallinga and Teunis approach (M1) and (B) susceptible-exposed-infectious-recovered (SEIR) model-based approach

## Conclusions

We examined the *R*_*t*_ of COVID-19 to monitor the impact of control measures on disease transmissibility. Taking transmission arising from imported cases into account, disease transmissibility of COVID-19 reached its peak on 16 January 2020 before Zhejiang declared the highest-level public health emergency and the government imposed a lockdown in Wuhan. As suggested by other investigations, approximately 35% of cities in China have ≥50% chance of having a COVID-19 case imported from Wuhan within 3 weeks before the lockdown *(12)*. A fast growth in disease transmission during the initial phase of epidemic can thus be expected, especially in areas that have tight travel connections with the source region *(13)*. Since *R*_*t*_ would surpass unity upon the emergence of only a few imported cases, COVID-19 was able to sustain in Zhejiang during the initial phase of the epidemic. Therefore, early preparation is strongly recommended to countries with limited imported cases at the moment.

We showed the transmission of COVID-19 reduced and remained at a low level not only after the lockdown of Wuhan, but also after the implementation of strict social-distancing measures in Zhejiang. As a result, disease transmission in the population was interrupted (*R*_*t*_<1) within several weeks alongside a drop in local cases. The decline in disease transmissibility might be attributed to the early declaration of public health emergency in Zhejiang, followed by implementation of large-scale lockdowns which seemed to interrupt and slow down the transmission of COVID-19 in the population. Our findings could thus inform high-risk regions on disease control planning in a timely manner.

One key limitation of our analysis is that the assumption of serial interval in the estimation is based on earlier estimates from Li *et al. (11)*. The estimates of *R*_*t*_ shall be improved if more updated knowledge of the pathogen are available.

## Data Availability

Data are available from the authors upon reasonable request and with permission of the Zhejiang Provincial Center for Disease Control and Prevention.

## Acknowledgments

We thank the physicians and staffs at Hangzhou, Huzhou, Jiaxing Wenzhou, Shaoxing, Ningbo, Quzhou, Jinhua, Zhoushan, Lishui, Taizhou Municipal Center for Disease Control and Prevention for their support and assistance with this investigation. The work is supported by National Natural Science Foundation of China (31871340, 71974165).

## Ethics

As this investigation was a part of the surveillance of emerging infectious diseases of Zhejiang Provincial Center for Disease Control and Prevention (CDC), ethics approval and informed consent were exempted by CDC. The data were analyzed anonymously.

## Author Bio

Dr. Chong is an Assistant Professor in the School of Public Health and Primary Care, The Chinese University of Hong Kong. He is also a Biostatistician in the Centre for Clinical Research and Biostatistics, Faculty of Medicine, The Chinese University of Hong Kong. His primary research interests are infectious disease epidemiology and environmental health.

